# Vaccination for some childhood diseases may impact the outcome of covid-19 infections

**DOI:** 10.1101/2020.09.02.20186528

**Authors:** Irene Gobe, Garesego F Koto, Margaret Mokomane, Kesaobaka Molebatsi, Ishmael Kasvosve, Modisa S Motswaledi

## Abstract

**Background:** COVID-19 found the world in a state of unpreparedness. While research efforts to develop a vaccine are on-going, others have suggested the use of available vaccines to boost innate immunity.

**Objective:** We analysed three databases: UNICEF Immunization Coverage, Worldometer Corona Virus Updates and World Bank List of Economies to establish the association, if any, between vaccination for various diseases and COVID-19 death rates and recoveries across world economies.

**Results:** Mean percentage death rates were lower in countries that vaccinated for Hepatitis-B birth dose (2.53% vs 3.79%, p = 0.001), Bacille Calmette-Guérin Vaccine (2.93% vs 5.10%, p = 0.025) and Inactivated Polio Vaccine 1^st^ dose (2.8% vs 4.01%, p = 0.022) than those which did not report vaccination. In high income countries, a significant negative correlation with death rates was observed with vaccination for Measles-containing vaccine 2^nd^ dose (r = –0.290, p = 0.032), Rubella-containing vaccine 1^st^ dose (r = –0.325, p = 0.015), Hepatitis B 3^rd^ dose (r = –0.562, p = 3.3 x10^−5^), Inactivated Polio vaccine 1^st^ dose (r = –0.720, p = 0.008). Inactivated Polio Vaccine 1st dose and Measles-containing vaccine 2^nd^ dose also correlated with better recoveries. In Low Income countries, only Rubella-containing vaccine correlated with lower deaths while Yellow fever vaccine was associated with poorer recoveries.

**Conclusion:** Our analysis corroborates the potential benefit of vaccination and warrant further research to explore the rationale for repurposing other vaccines to fight COVID-19.

## Introduction

A number of recent publications have raised conversations regarding the potential benefits of BCG vaccine in mitigating the adverse effects of SARS COV-2 infection(1, 2). Such hypotheses were inspired by past reports where BCG vaccination was observed to have benefits beyond prevention of tuberculosis alone. These included protection against some viral infections(3) as well as overall reduction in childhood mortality occasioned by other infectious agents(4). A recent publication based on country policies reported that countries with a long history of BCG vaccination experienced lower numbers of SARS COV-2 infections and fewer deaths(1). Another study found a strong correlation between the BCG index and reduced COVID-19 mortalities in European countries (5).

While most vaccines confer immunity through antigen-specific immune responses, some reports suggest that vaccination against one disease may confer protection against another, as evidenced in the upregulation of a wide range of innate immune responses to BCG vaccine(6). This phenomenon is attributed to the development of “trained” immunity, in which BCG-activated monocytes are primed to produce more cytokines such as IL-1β, IL-6, IFN-γ and TNF(2).

In fact, other investigators have reported increased expression of pathogen-associated molecular patterns on cells of monocytic origin up to one year after exposure to BCG, enabling a more robust induction of TH1/Th17 responses(7). BCG vaccination was also noted to enhance production of such cytokines as IFN-ƴ, IL-2 and IL-12, which are important for potentiation of cell mediated immunity(8). The above notwithstanding, other investigators have cast doubt on the potential benefits of BCG vaccination in the fight against COVID-19(9).

The afore-mentioned discussions have inevitably culminated on the debate to consider the potential prophylactic use of BCG against COVID-19(10), with some data corroborating the anti-COVID-19 effect(11). In the current report, we analysed global vaccination coverage and global COVID-19 data to determine if any association existed between the degree of vaccine coverage and the death rates attributed to COVID-19, as well as recovery rates.

## Methods

We studied global vaccination coverages as reported by UNICEF(12) and compared them to COVID-19 deaths and recoveries across the world as reported on the Worldometer(13). These vaccines included the measles-containing-vaccine first dose (MCV1), measles-containing vaccine second dose (MCV2), Pneumococcal Conjugate Vaccine – 3^rd^ Dose (PCV3), Rubella-containing vaccine first dose (RCV1), Rotavirus-containing vaccine, (RotaCV), Diphtheria and tetanus toxoid and pertussis-containing vaccine (DTP3), Hepatitis B vaccine – Birth dose (HepB_BD), Hepatitis B third dose (HepB3), *Haemophilus influenzae* type b-containing vaccine (HibV), Bacille Calmette-Guérin Vaccine (BCG), inactivated Polio virus vaccine (IPV) and Yellow fever vaccine (YFV).

The death rate for each country was computed as the total number of deaths per million divided by the total number of confirmed cases per million, expressed as a percentage. The independent samples t-test was used to compare the mean percentage of COVID-19 deaths between countries that reported vaccinations and those which did not, using data extracted on the 3^rd^ August 2020 from the COVID-19 Worldometer. Death and recovery data were available for 168 and 175 countries, respectively. However, some vaccination data was missing for some countries and this resulted in different sample sizes as reflected in the analyses. Mean vaccination coverages were correlated with recovery and death rates where such vaccination data was available. Otherwise we compared country death rates and recoveries according to categories of whether vaccine data was reported or not. Data was also disaggregated according to the World Bank economic classification (June 2020)(14).

## Results

The mean death rate (±sd) from COVID-19 for 168 countries was 3.2±3.5% with a range of 0.06–29.3%, while the mean recovery rate for 175 countries was 68.2±22.3%. Recoveries ranged from 8–100%. The results showing a comparison of mean COVID-19 death rates between vaccinating and non-vaccinating countries is shown in Table 1. Only Hepatitis B-BD, BCG and IPV (p = 0.01, p = 0.025 and p = 0.022, respectively) showed significant differences in mean COVID-19 death rates across vaccinating and non-vaccinating countries, with vaccinating countries showing significantly lower death rates. Other vaccines did not yield significant differences between vaccinating and non-vaccinating countries.

**Table 1:** Comparison of Covid-19 death rates in countries vaccinating or not vaccinating for certain diseases

| Vaccine name* | Vaccinating Countries |  | Non-Vaccinating Countries |  | Two-tailed significance |
| --- | --- | --- | --- | --- | --- |
|  | N | COVID-19 Death Rate (%±SD) | N | COVID-19 Death Rate (%) |  |
| MCV2 | 144 | 3.38±3.66 | 24 | 2.40±2.11 | 0.217 |
| PCV3 | 130 | 3.43±3.82 | 38 | 2.60±1.87 | 0.191 |
| RCV1 | 143 | 3.34±3.67 | 25 | 2.67±2.17 | 0.360 |
| RotaCV | 92 | 3.29±3.95 | 76 | 3.20±2.87 | 0.857 |
| HepB_BDV | 72 | 2.53±1.81 | 96 | 3.79±4.28 | 0.010 |
| HepB3V | 160 | 3.11±3.34 | 8 | 6.09±5.22 | 0.153 |
| BCG | 143 | 2.93±3.23 | 25 | 5.10±4.4 | 0.025 |
| YFV | 36 | 2.88±2.06 | 132 | 3.35±3.35 | 0.478 |
| IPV1 | 113 | 2.81±3.38 | 55 | 4.1±3.56 | 0.022 |

\* MCV2: Measles containing vaccine 2<sup>nd</sup> dose, PCV3: Pneumococcal Conjugate vaccine 3<sup>rd</sup> dose, RCV1: Rubella-containing vaccine 1<sup>st</sup> dose, RotaCV: Rotavirus-containing vaccine, HepB-BD: Hepatitis B birth dose, HepB3V: Hepatitis B 3<sup>rd</sup> dose, BCG: Bacille Calmette-Guérin Vaccine, YFV: Yellow fever vaccine, IPV1: Inactivated Polio vaccine 1<sup>st</sup> dose.

Correlation of the aggregated data showed that the following vaccines were weakly associated with lower COVID-19 death rates: Rubella-containing virus (R = –0.191, p = 0.022), Hepatitis B3 (r = –0.201, p = 0.011) and BCG (r = –0.221, p = 0.008). None of the vaccines were associated with increased COVID-19-related death. The correlation between the number of years that any country was vaccinating for the diseases and death rates was only significant for IPV1 in high-income countries (r = –0.620, p = 0.042).

We observed that the percentage of recoveries was higher in non-vaccinating countries for the diseases shown in Table 2.

**Table 2:** Vaccines that show significant differences in mean percent Covid-19 recoveries.

| **Vaccine\*** | **No. countries** | **Recovery** | **No. countries not** | **Recovery** | **Two-tailed** |
| --- | --- | --- | --- | --- | --- |
| **Name** | **Vaccinating** | **%±SD** | **Vaccinating** | **%±SD** | **significance** |
| PCV3 | 131 | 65.6±22.7 | 44 | 76.0±18.9 | 0.007 |
| RotaC | 93 | 63.8±23 | 82 | 73.1±18.9 | 0.005 |
| HepB3 | 168 | 67.4±22.3 | 7 | 86.1±9.7 | 0.001 |
| YFV | 36 | 61.7±20.4 | 139 | 69.9±22.5 | 0.050 |

\*PCV3: Pneumococcal Conjugate vaccine, RotaC: Rotavirus-containing vaccine, HepB3: Hepatitis B 3<sup>rd</sup> dose, YFV: Yellow Fever vaccine

We also compared percent death and recovery rates according to income level of the countries. The results, summarized in Table 3, show a clear correlation of some vaccines coverages with lower death rates. Only Pol-3 correlated with a significant unfavourable outcome in high income countries. RCV-1 showed similar favourable outcomes in both high- and medium-income countries. IPV strongly correlated with lower death rates in high income countries and higher recoveries in upper middle-income countries, while YFV showed negative correlation with percent recovery.

**Table 3:**
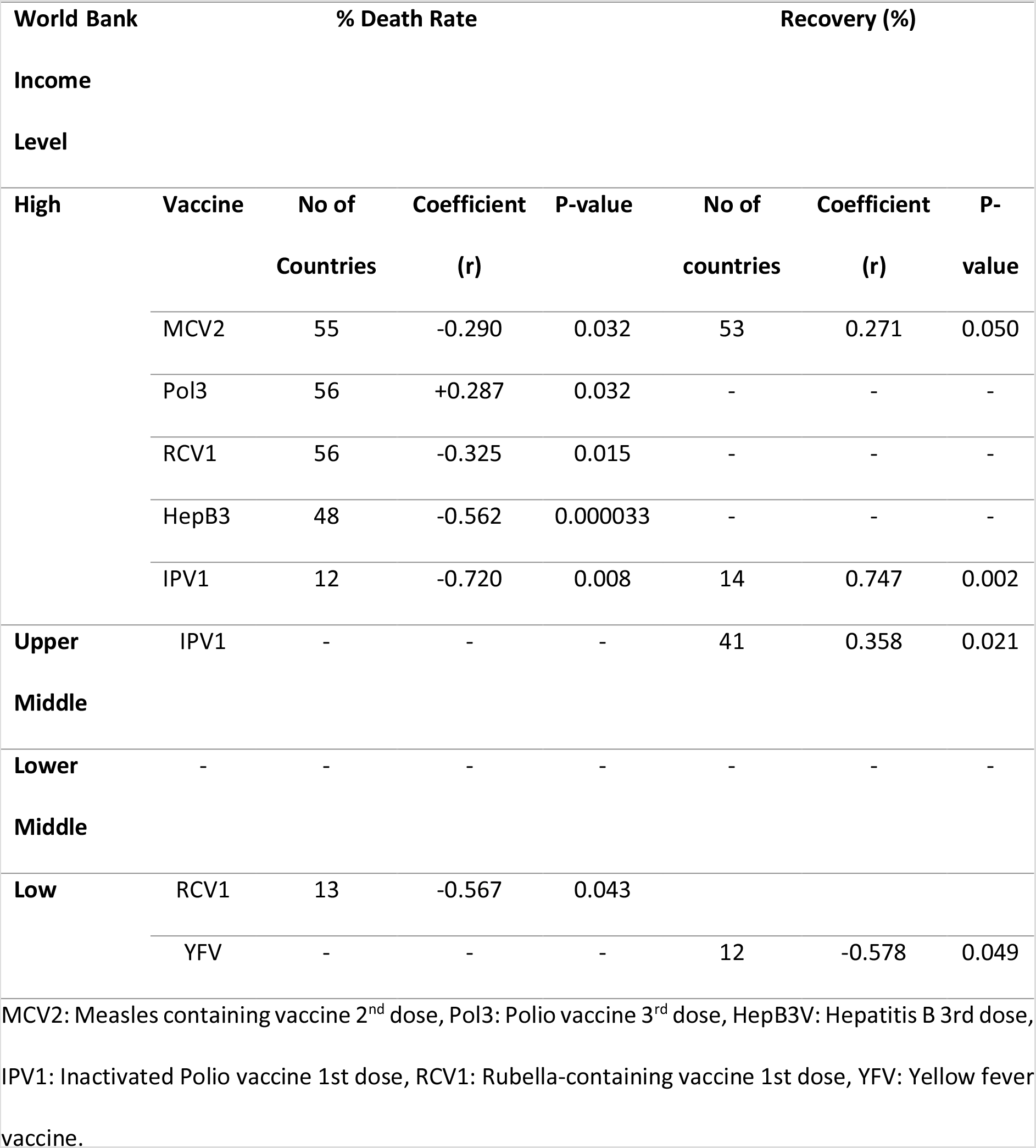
Correlation of average vaccine coverages with death and recovery rates according to World Bank Income-based classification

## Discussion

We undertook an analysis of COVID-19 deaths and recoveries in light of immunization with various vaccines to determine the association, if any, of vaccination in mitigating the adverse outcomes of infection with the SARS CoV-2 virus. Vaccination with Hepatitis B birth dose, BCG and inactivated Polio vaccine were associated with lower Covid-19 deaths. This is consistent with other reports where BCG vaccination was associated with lower mortalities and severity of the disease(1, 5). We also observed a weak but significant correlation suggesting that COVID-19 deaths are generally lower in countries where vaccination for BCG, Rubella-containing vaccines and Hepatitis B 3^rd^ dose are given. Moreover, we observed that vaccine coverage, rather than the longevity of use, has a better correlation with percent deaths attributed to COVID-19.

We note that death or recovery is also a function of the effectiveness of the health care system in any country. We presumed that higher income countries would generally afford better health care systems and therefore better recoveries and lower deaths.

Paradoxically, percent recoveries were significantly higher in non-vaccinating countries for Pneumococcal Conjugate vaccine (PCV), RotaCV and Hepatitis B-3 vaccines. We hypothesize that if vaccines, as has been established, promote innate immunity (4, 7, 15, 16), then both death rates and recoveries should be expected to be favourable in vaccinated individuals. However, since recovery includes individuals who must have been hospitalized for symptomatic presentation or those who tested positive but never needed to recover since they were asymptomatic, those with competent immune systems would not be expected to be hospitalized, and would therefore be underrepresented among documented recoveries. We propose that this under-representation may account for the higher recoveries in non-vaccinating countries for the specified vaccines.

There is evidence suggesting that vaccination against tuberculosis, and other vaccinations presented in this work, could provide protection or ameliorate the severity of COVID-19 deaths. Given the gravity of the current COVID-19 threat and the relative unpreparedness of most developing countries, we strongly recommend coordinated clinical trials on vaccination or revaccination as an additional strategy to mitigate the effects of COVID-19. This may imply carrying out clinical trials in designated areas, especially in countries such as Botswana, where the pandemic is still evolving.

## Study limitations

Our study has several limitations. There could be confounding factors in our data that affect both vaccination coverage and COVID-19 deaths such as genetic diversity, age of population, prevalence of comorbidities, global traveling, population density and quality of health care services. While we did not test for causal effects of vaccination coverage on COVID-19 death rates, we observe that our findings corroborate earlier reports on the potential of other vaccines in mitigating the deleterious effects of COVID-19.

Another potential limitation is the fact that the disease is still developing in many countries, so the true resilience of many countries is still to be tested. It is also not clear how to interpret lower recoveries especially in view of the different definitions of recovery. In Botswana, for example, recovery was initially defined in the context of two successive nucleic acid tests at least twenty four hours apart. The current definition now relies on the clinical manifestations and age of the infection. Nevertheless, the data seems to corroborate reports alluding to potential benefit of BCG of some vaccines against COVID-19.

## Data Availability

All data is available upon request

## Data availability

The data used in this analysis is available as a public resource from UNICEF, World Bank and Worldometer databases.

